# Phenotypic and genetic characterization of different modes of lifetime nicotine use in the All of Us Research Program

**DOI:** 10.64898/2026.09.11.26361370

**Authors:** Feiyang Huang, Pamela N Romero Villela, Zhen Luo, Alex P Miller, Pamela Madden, Arpana Agrawal, Alexander S Hatoum, Emma C Johnson

## Abstract

**Objective:** Existing genetic studies of nicotine consumption have largely focused on cigarette smoking, despite other modes emerging. Different nicotine use modes have varying use patterns and user characteristics that may contribute to distinct environmental and genetic influences. This study aims to identify the shared and specific genetic risks for five nicotine use modes.

**Methods:** We performed a common variant multi-ancestral genome-wide association study (cross-ancestry N = 304,213) of five types of nicotine use in the *All of Us* Research Program: lifetime use of electronic nicotine (e-nicotine) products, cigars, tobacco in a hookah, smokeless tobacco, or at least 100 cigarettes.

**Results:** Nicotine users, regardless of mode, were more likely to be male and of younger age (except for cigarette use), and all modes of use were positively associated with tobacco use disorder diagnoses. GWAS meta-analyses of cigarette, e-nicotine, cigar, and smokeless tobacco use identified 87, 4, 1, and 1 genome-wide risk loci, respectively (none for hookah), including previously identified genes (e.g., *CHRNA4* for cigarettes). Cigarette, e-nicotine, and smokeless tobacco use showed strong genetic correlations (r_g_=0.82-0.92), while cigar and hookah use were strongly genetically correlated (r_g_=0.87). Cross-trait correlations diverged: cigarette, e-nicotine, and smokeless tobacco use showed the strongest positive correlations with psychiatric disorders, negative urgency, and neuroticism, whereas cigar and hookah use showed null-to-weak correlations with psychiatric disorders but positive correlations with sensation-seeking and openness.

**Conclusions:** Our findings reveal important phenotypic and genetic distinctions across modes of nicotine use, particularly in relation to personality traits such as impulsivity and to socioeconomic context.

## INTRODUCTION

Nicotine use is a major behavioral risk factor^1^ that contributes to preventable diseases (e.g., cancer, lung disease) and other physical and mental health problems^2^. Although cigarette smoking has been the dominant form of nicotine consumption and the primary focus of research, multiple non-cigarette nicotine products are increasingly used. Among these, electronic nicotine (e-nicotine) products represent one of the most rapidly increasing modes of nicotine use, becoming the second most frequently reported nicotine product after cigarettes, with prevalence among adults in the United States rising from 4.5% in 2019 to 7.0% in 2024^3,4^. Similarly, cigar sales increased by $0.8 billion between 2009 and 2020^5^.

These nicotine products differ in their patterns of use and exposure contexts. For example, hookah smoking often occurs in group settings, in urban environments near universities, and sessions may last for longer periods^6,7^, while cigar smoking is typically characterized as an occasional activity associated with higher socioeconomic status and social practices^8^. In contrast, cigarette smoking is more often related to nicotine dependence and stress coping^8^. These differences in contexts and product characteristics suggest that the initiation and use of various nicotine products may occur through distinct behavioral pathways.

Genome-wide association studies (GWAS) have identified numerous loci associated with nicotine dependence and smoking initiation. These studies have also characterized genetic correlations between nicotine use behaviors and other health-related traits, generally finding positive correlations between nicotine use and other substance use, cardiovascular and respiratory disease, and psychiatric disorders (e.g., ADHD). However, the majority of genetic studies of nicotine use have focused exclusively on cigarette smoking, leaving other commonly used nicotine products relatively understudied^9,10,11^. While twin studies have demonstrated a significant genetic correlation between different nicotine products^11^, the extent to which different modes of nicotine use share genetic factors remains unclear, as does the consistency of their genetic associations with other health outcomes.

In this study, we analyzed five types of nicotine use in the *All of Us* (AoU) Research Program^12^: whether participants had ever used e-nicotine products, smoked a cigar, smoked tobacco in a hookah, used smokeless tobacco products, or smoked at least 100 cigarettes. We first conducted phenotypic analyses to examine how these phenotypes differ by sex and age, and evaluated their associations with tobacco use disorder. Then, we performed GWAS in three groups of participants who were most genetically similar to people of European ancestry (N = 181,797), Admixed American ancestry (N = 58,110), and African ancestry (N = 64,306; see **Table 1**), followed by a cross-ancestry meta-analysis (N = 304,213, Neff_max_= 174,840, Neff_min_= 12,579). In addition to comparing the genetic correlations among the five modes of nicotine use, we also studied how genetic correlations with psychiatric disorders, personality, and other substance use traits compared across nicotine modalities.

**Table 1.** Definitions of nicotine-use phenotypes based on survey questions, with descriptive statistics for sex, age, and sample size across three ancestry groups.

| Phenotype | Exact question in the survey |  |  |  |  |  |  |  |
| --- | --- | --- | --- | --- | --- | --- | --- | --- |
| Cigar ever-use | Have you ever smoked a traditional cigar, cigarillo, or filtered cigar, even one or two puffs? |  |  |  |  |  |  |  |
| ≥ 100 Cigarette smoking | Have you smoked at least 100 cigarettes in your entire life? (There are 20 cigarettes in a pack.)? |  |  |  |  |  |  |  |
| E-nicotine ever-use | Have you ever used an electronic nicotine product, even one or two times? (Electronic nicotine products include e- cigarettes, vape pens, hookah pens, personal vaporizers and mods, e-cigars, e-pipes, and e-hookahs.) |  |  |  |  |  |  |  |
| Hookah ever-use | Have you ever smoked tobacco in a hookah, even one or two puffs? |  |  |  |  |  |  |  |
| Smokeless ever-use | Have you ever used smokeless tobacco products, even one or two times? (Smokeless tobacco products include snus pouches, Skoal Bandits, loose snus, moist snuff, dip, spit, and chewing tobacco.) |  |  |  |  |  |  |  |
| Phenotype | Male<sub>users</sub> | Male<sub>nonusers</sub> | Mean Age<br>(Nonuser - User) | 95% CI ΔAge | Ancestry | Case | Control | N<sub>effective</sub> |
| Cigar ever-use | 57% | 26.6% | 55.0 - 51.6 | [3.25,3.5] | AFR | 18,570 | 42,953 | 51,859 |
|  |  |  |  |  | AMR | 15,696 | 40,951 | 45,388 |
|  |  |  |  |  | EUR | 77,140 | 100,861 | 174,840 |
| ≥ 100 Cigarette smoking | 46.2% | 32.3% | 51.9 - 56.3 | [-4.47,-4.23] | AFR | 28,867 | 32,961 | 61,557 |
|  |  |  |  |  | AMR | 17,195 | 39,220 | 47,816 |
|  |  |  |  |  | EUR | 75,632 | 102,126 | 173,809 |
| E-nicotine ever-use | 40.2% | 37.7% | 56.8 - 42.2 | [14.42,14.73] | AFR | 9,514 | 52,202 | 32,189 |
|  |  |  |  |  | AMR | 10,112 | 46,693 | 33,248 |
|  |  |  |  |  | EUR | 28,212 | 150,657 | 95,049 |
| Hookah ever-use | 40.9% | 37.5% | 57.7 - 41.4 | [16.07,16.36] | AFR | 9,837 | 51,693 | 33,057 |
|  |  |  |  |  | AMR | 11,457 | 45,326 | 36,581 |
|  |  |  |  |  | EUR | 28,594 | 149,566 | 96,019 |
| Smokeless ever-use | 75.6% | 34.3% | 54.5 - 46.8 | [7.48,7.9] | AFR | 3,347 | 58,428 | 12,663 |
|  |  |  |  |  | AMR | 3,341 | 53,573 | 12,579 |
|  |  |  |  |  | EUR | 21,158 | 157,978 | 74,636 |

## METHODS

### Sample

This study utilized data from the controlled-tier version 8 release of the *All of Us* Research Program, a cross-sectional, large-scale biobank of individuals aged 18 and older residing in the United States ^12^.

All participants with available short-read whole genome sequencing (srWGS) data and responses for the “Lifestyle” survey were considered for analysis (N = 304,213). Detailed case– control sample counts by phenotype and ancestry are provided in **Table 1**.

### Phenotype definition

We used data from the Lifestyle survey to create separate binary variables of self-reported lifetime use of five different types of nicotine products (1=“yes”, 0=“no”). Specifically, participants were asked whether they had ever used e-nicotine products, cigars, tobacco in hookahs, smokeless tobacco products, or smoked at least 100 cigarettes in their lifetime (exact questions and sample sizes provided in **Table 1**). Participants with a response of “no” were classified as controls for the corresponding nicotine product.

### Phenotypic analyses

We tested whether endorsement of the different nicotine use phenotypes differed by sex and age, controlling for self-reported race as a covariate. We also compared associations with a lifetime tobacco use disorder (TUD) diagnosis across the different nicotine use phenotypes for participants with available EHR data, using a series of separate logistic regression models adjusting for sex, age, and self-reported race, as well as a multivariable model including all phenotypes simultaneously to assess their independent associations with TUD.

### Genotype quality control

Whole genome sequence data were initially quality controlled by the AoU research team using Illumina’s DRAGEN pipeline^13^. For these analyses, we additionally filtered by minor allele frequency >1% to only include common single-nucleotide polymorphisms (SNPs) and insertions/deletions in our analyses.

### Genome-wide association analyses and Multi-ancestry meta-analysis

Genome-wide association analyses were performed within the three largest genetic ancestral population subsets as defined by the AoU research team: European (EUR), Admixed American (AMR), and African (AFR) ancestry subgroups (see **Table 1** for sample sizes). We used PLINK2^14^ to perform logistic regression analyses for each tobacco use outcome, covarying for age in general, sex, and the first ten within-ancestry genetic principal components. The resulting summary statistics for each ancestral group were meta-analyzed using MR-MEGA v.0.2 to account for heterogeneity correlated with genetic ancestry, thereby improving statistical power^15^. MR-MEGA constrains the number of genetic axes (T) to be at least two fewer than the number of included ancestries (K). Therefore, with three ancestry groups (K = 3), we used T = 0, which provides a meta-analysis framework but does not model heterogeneity as a function of genetic ancestry.

### FUMA annotation and gene-based analyses

First, we lifted over the GWAS summary statistics from hg38 to hg37 using GWASLab^16^ v. 3.6.3 on Python v. 3.9.12 before conducting follow-up analyses. We used FUMA^17^ version 1.8.2 to define “independent significant SNPs” as those that reached genome-wide significance (*p* < 5e-8) and were independent of each other at r^2^ < 0.6, and “lead SNPs” as those SNPs that were independent of each other at r^2^ < 0.1. To enable interpretation of multiple correlated SNPs as a single association signal, “genomic risk loci” were defined by merging linkage disequilibrium (LD) blocks of independent significant SNPs within a 250 kb distance. We used the appropriate ancestry-matched 1000 Genomes Phase 3 sample as the LD reference panel. Gene-based analyses were conducted using MAGMA via FUMA, which aggregates single SNPs to the gene level and tests the joint association of all markers within a gene with the phenotype.

### Cross-ancestry genetic correlation analyses

We used POPCORN^18^ version 1.1 to estimate cross-ancestry genetic correlations for the five different nicotine use phenotypes, performing pairwise comparisons of each phenotype across EUR, AFR, and AMR populations. POPCORN provides two different estimates of the shared genetic basis of a trait between ancestral populations. Specifically, genetic effect correlation (r_ge_) assesses the degree to which the same variants cause phenotypic variance in both populations, regardless of allele frequency, whereas genetic impact correlation (r_gi_) weights common alleles more heavily than rare ones to accurately reflect their true contribution to phenotypic variance within each specific population. In our analysis, we estimated both r_ge_ and r_gi_^18^.

### SNP-heritability and genetic correlation analyses within nicotine use phenotypes

We used linkage disequilibrium score regression (LDSC) to estimate the SNP-heritability of the nicotine use phenotypes in the EUR and AFR ancestry subsamples^19^. As the LDSC *z*-scores for the AFR subsample were below the recommended threshold of 4^20^, indicating inadequate power for genetic correlation estimation, genetic correlation analyses were performed only in the EUR subsample.

### Genomic Structural Equation Modeling (GenomicSEM)

We applied Genomic Structural Equation Modelling (Genomic SEM) using the GenomicSEM R package (version 0.0.5c)^21^ to model the genetic relationships between the five nicotine use phenotypes in the EUR ancestry subsamples. First, we constructed a common factor model, so the five nicotine use phenotypes loaded on one common factor. Considering the results of our pair-wise genetic correlation analyses, we also tested a two-factor model, where cigarette, e-nicotine, and smokeless tobacco use loaded on one factor, and hookah and cigar use loaded on a second factor. For both models, indicators were allowed to load freely while the variances of any latent factors were scaled to 1.0, and we used weighted least squares estimation. Model fit was evaluated using the Chi-square test, the comparative fit index (CFI), with CFI > .9 deemed as an acceptable fit^22^, and the Standardized Root Mean Square Residual (SRMR), with SRMR < 0.08 considered as a good fit, as seen in past GenomicSEM models. To test the difference in fit between the single and two-factor model, we compared the goodness of fit between the two models using a chi-square difference test.

### Genetic correlation analyses with other phenotypes

We also used LDSC to estimate the genetic correlations between the different nicotine use phenotypes in European ancestry, and between the nicotine use phenotypes and 30 other traits, including other substance use behaviors and disorders, psychiatric disorders, physical health, personality traits, cognition, and socioeconomic status-related phenotypes (see **ST1**).

## RESULTS

### Phenotypic results

Across all ancestry groups, smoking ≥100 lifetime cigarettes was the most commonly endorsed behavior, followed by cigar and hookah use (sample sizes provided in **Table 1** and shown in **Figure 1A**). Exclusive use of smokeless tobacco was relatively rare (N_AFR_=243, N_AMR_=159, N_EUR_=1120), accounting for <10% of individuals reporting any smokeless tobacco use (including co-use with other nicotine products) across all ancestry groups. The most prevalent co-use of nicotine products across all ancestry groups was cigarette smoking and cigar use, representing approximately 31% of all cigar users in AFR, 18% in AMR, and 25% in EUR (**Figure 1A**).

**Figure 1.**
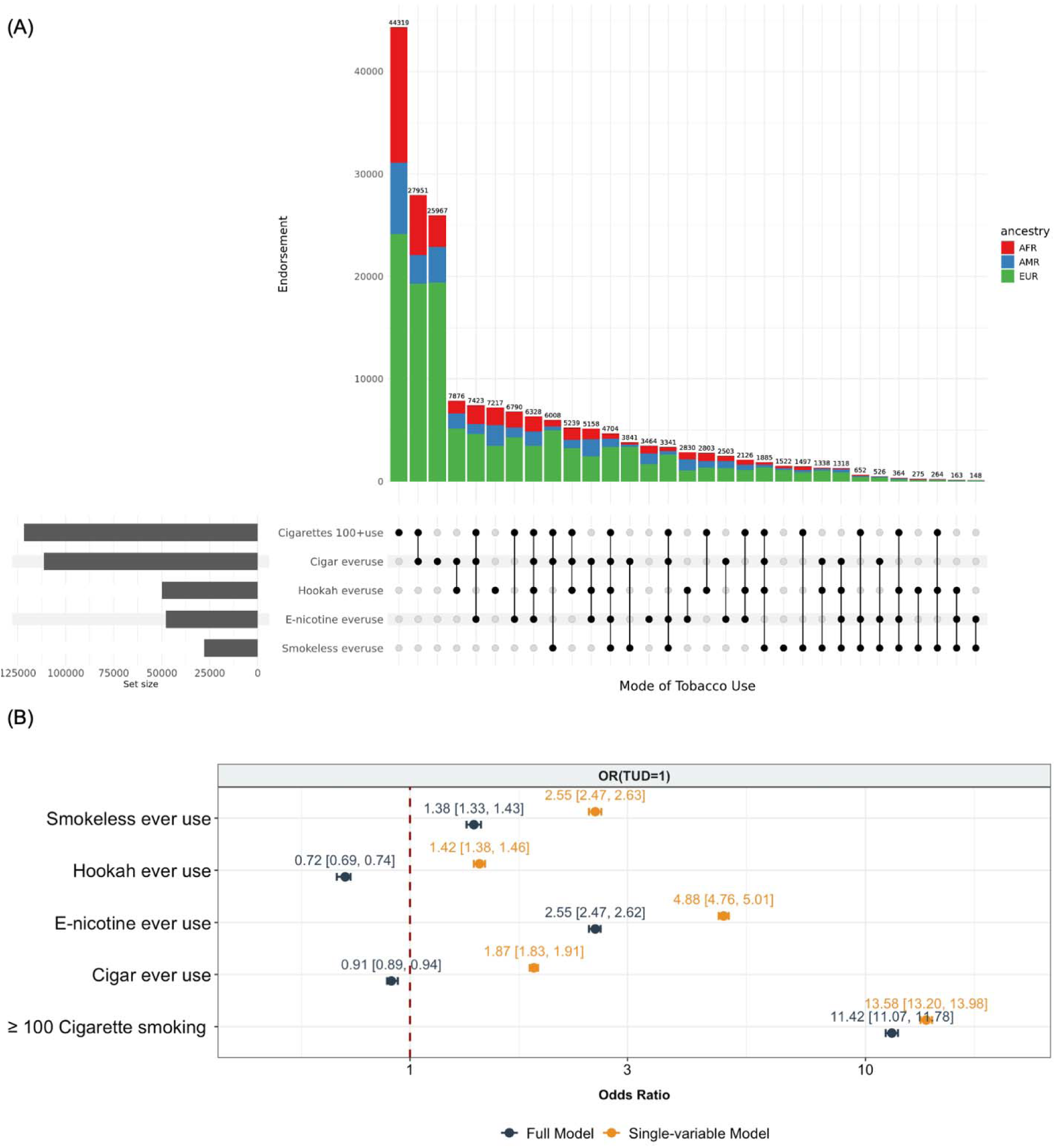
Phenotypic overlap and association models across five nicotine use modes. (A) UpSet plot illustrating the co-use patterns of five nicotine use phenotypes. The intersection bars are colored by ancestry. Total counts for each unique combination of product use are displayed above the corresponding bars. (B) Single-variable (orange) vs. full multivariable (dark slate) model comparisons showcasing OR and 95% CI for being diagnosed as TUD. Red dashed vertical lines in (B) indicate the OR = 1.0. Point estimates are explicitly labeled alongside their respective 95% intervals.

Participants endorsing lifetime use were significantly younger than non-users for all nicotine use phenotypes except ≥100 cigarette smoking, perhaps because of the longer duration necessary to meet the ≥100 cigarette criterion (Mean_use_ = 56.27, Mean_non-use_= 51.92; **ST2**). In particular, those endorsing lifetime use of hookah (Mean_use_ = 41.52, Mean_non-use_= 57.74) and e-nicotine (Mean_use_ = 42.24, Mean_non-use_= 56.82) were considerably younger than non-users. After adjusting for age and self-reported race, males were significantly more likely than females to endorse all nicotine phenotypes, with the strongest male predominance observed for smokeless tobacco (OR =7.23, 95% CI=[7.02, 7.45]; **Table 1, ST2**).

In single-phenotype models, all nicotine use behaviors were significantly positively associated with lifetime TUD diagnosis (ORs = 1.42 (hookah use) - 13.58 (≥100 cigarettes)). However, in the multivariable model adjusting for all modes of nicotine use, the associations with TUD for hookah and cigar use became negative (hookah OR = 0.72, 95% CI = [0.69, 0.74]; cigar OR = 0.91, 95% CI = [0.89, 0.94]) and remained statistically significant. The remaining three nicotine use behaviors remained significantly positively associated with TUD, although effect sizes were attenuated compared to the single-phenotype models (**Figure 1B, ST2**).

### Genome-wide association study results

Our results showed a highly polygenic burden for ≥100 lifetime cigarette use, for which the cross-ancestry meta-analysis identified 87 genome-wide significant loci (*p* < 5 × 10⁻) and 113 significant genes in MAGMA^23^ gene-based tests. This signal was also evident in the EUR subsample, where 35 independent genome-wide significant loci (25 overlapped with meta-analysis) and 74 MAGMA-significant genes were identified (51 overlapped with meta-analysis), including *BDNF,* which was previously identified in a GWAS of TUD^24^. In the AFR subsample, there were 3 genome-wide risk loci and 6 MAGMA-significant genes identified for this trait (5 overlapped with the meta-analysis: *C10orf2*, *FAM178A*, *LZTS2*, *MRPL43*, *SEMA4G*). E-nicotine use also showed notable signals, with 4 genome-wide significant loci and 8 MAGMA-significant genes in the meta-analysis; the EUR ancestry GWAS similarly identified 3 genome-wide significant loci and 11 MAGMA-significant genes (4 overlapped with meta-analysis: *FBXL16*, *NCAM1*, *SHQ1*, *WDR24*). In contrast, meta-analyses of cigar and smokeless tobacco use each only yielded 1 genome-wide significant locus, and no genome-wide significant SNPs were identified for hookah use (**Figure 2**, **ST3-6**).

**Figure 2.**
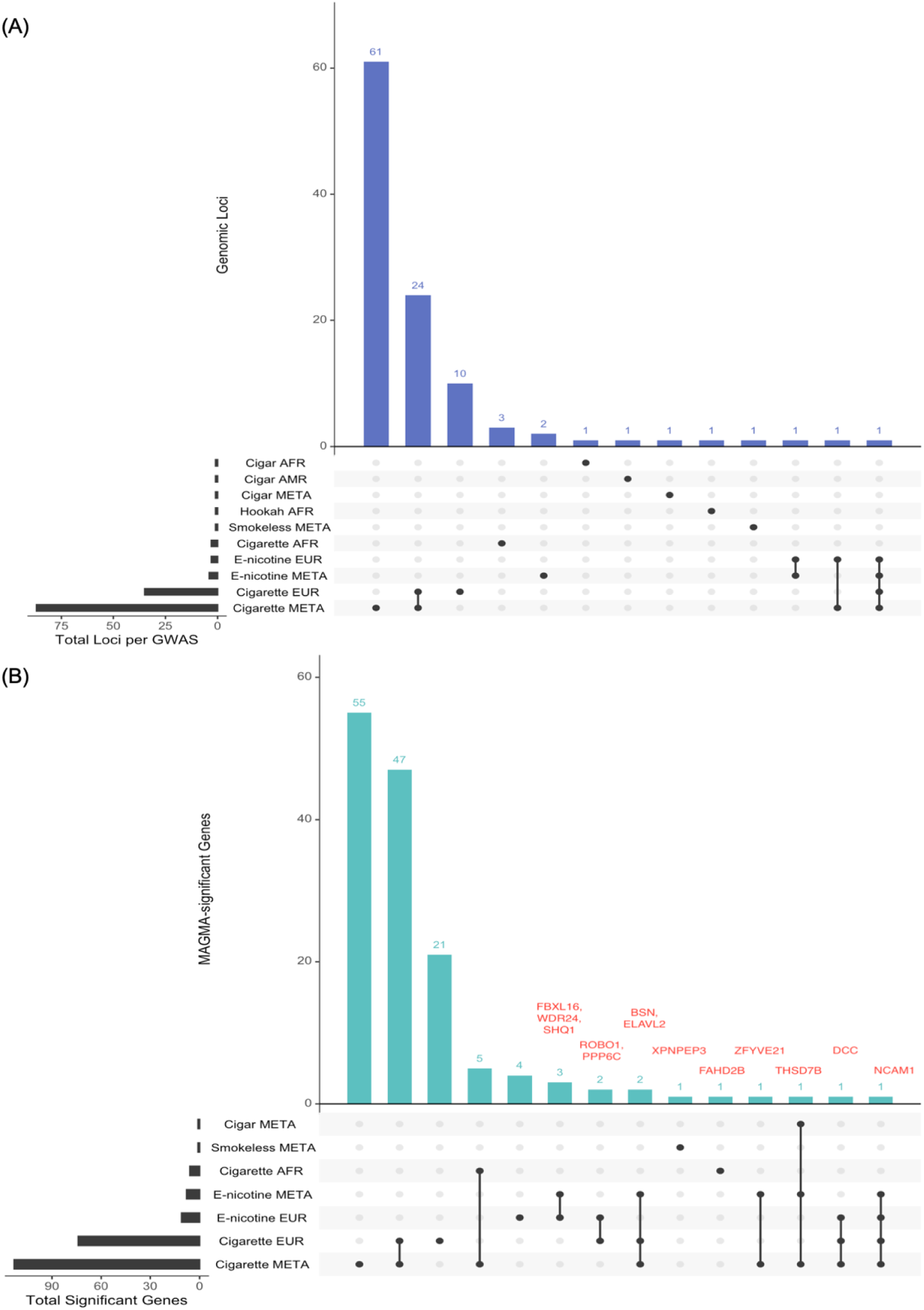
Shared genomic loci and MAGMA-significant genes across nicotine use GWAS. (A) UpSet plot illustrates the overlap of GWAS loci across ancestral groups and meta-analysis. Overlapping loci were defined by their physical start and end positions. Horizontal bars show the total number of independent genomic loci discovered per GWAS, vertical intersection bars represent the number of genomic regions shared across GWAS. (B) UpSet plot shows the overlapped matches of genome-wide significant genes via MAGMA (*p*_bonferroni_< 0.05). The horizontal axis represents the total number of unique significant genes per GWAS, the vertical bars indicate genes shared across GWAS. For sets with 3 or fewer, gene names are provided.

No genome-wide significant SNPs were identified for hookah, cigar, or smokeless tobacco use within the EUR subsample. In the AFR subsample, 1 genome-wide risk locus was identified for cigar use (lead SNP:rs784329), and 1 observed for hookah use (lead SNP:rs149965463). In AMR, 1 genome-wide risk locus was identified for cigar use (lead SNP:rs12358362).

Cross-trait gene-based results suggested shared genetic architecture across nicotine-use phenotypes, with *THSD7B* being significant in the cross-ancestry meta-analyses of ≥100 cigarette, e-nicotine, and cigar use, and *NCAM1* implicated in both ≥100 cigarette and e-nicotine use (see **ST6**). Overlaps with GWAS Catalog further suggested that identified loci are correlated not only to smoking-related traits: across analyses, 77 independent genome-wide significant loci overlapped with previously reported smoking-related associations, 81 overlapped with psychiatric or psychological traits, and 34 overlapped with non-tobacco substance-use traits, including alcohol, cannabis, and opioid use (see **ST7**).

### Genetic correlations between nicotine use phenotypes

Cross-population genetic correlations estimated from POPCORN showed relatively large standard errors across ancestry pairs, particularly for hookah and smokeless tobacco use ≥1.00. Cigarette use exhibited consistently high genetic correlations across all ancestry pairs, from AFR–AMR (r_gi_=0.65, SE=0.076) to AMR–EUR (r_gi_=0.84, SE=0.068). For e-nicotine use, significant genetic correlations were observed for AFR–EUR (r_gi_=0.49, SE=0.197) and AMR– EUR (r_gi_=0.68, SE=0.098). Cigar use showed significant correlations for AFR–AMR (r_gi_=0.65, SE=0.290) and AMR–EUR (r_gi_=0.72, SE=0.146; **Figure 3A, ST8**).

**Figure 3.**
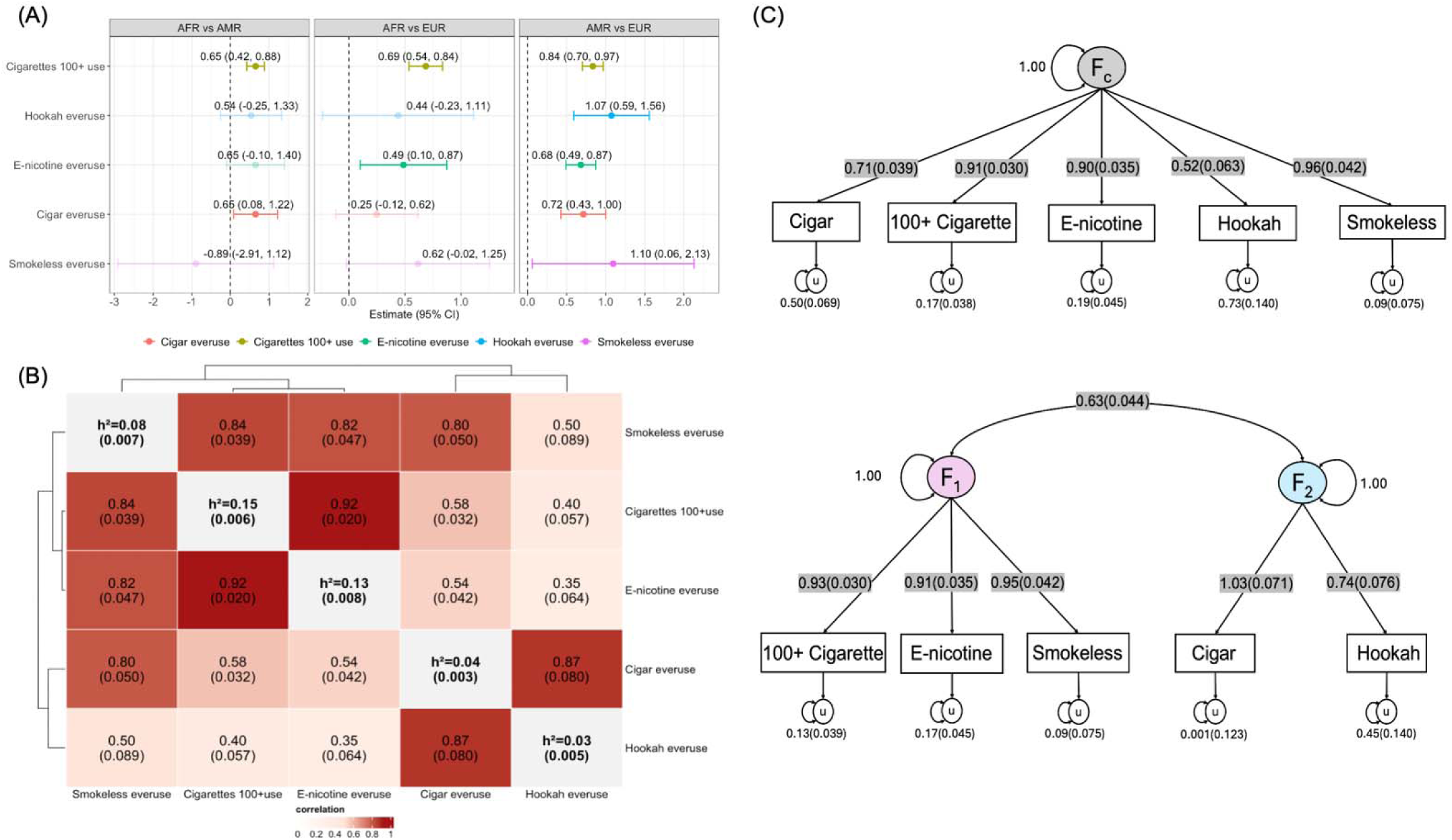
Cross-ancestry genetic correlations (rgi) for each nicotine use phenotype(panel A), genetic correlations between all five nicotine use phenotypes within EUR ancestry samples (panel B), and results of genomic SEM within EUR ancestry samples (panel C). Heritabilities are shown on the diagonal in panel B.

Within-ancestry genetic correlation analyses using LDSC were only conducted in the EUR ancestry group, as heritability estimates in the AFR group were insufficiently powered for most phenotypes (see **ST9**). All five nicotine use phenotypes were significantly genetically correlated, although the extent of the correlations varied substantially (r_g_s from 0.35 to 0.92, **Figure 3B**). The strongest genetic correlation was observed between ≥100 cigarettes and e-nicotine use (r_g_ = 0.92, SE = 0.020). Although hookah use was strongly genetically correlated with cigar use (r_g_ = 0.87, SE = 0.080), it showed much smaller genetic correlations with the other modes (r_g_ = 0.35 to 0.58). Smokeless tobacco use showed genetic correlations >0.8 with every other phenotype except for hookah use (r_g_ = 0.50, SE = 0.089).

In the GenomicSEM common factor model, ≥100 cigarettes, e-nicotine, and smokeless tobacco use showed very strong loadings on the latent factor (all standardized loadings > 0.9), whereas cigar and hookah use loaded slightly lower (0.71 and 0.52, respectively; **Figure 3C**). This model had a relatively poor fit (χ² = 144.1 (df = 5), p = 2.3963e-29; AIC = 164.1; SRMR = 0.15; CFI = 0.96). In contrast, the two-factor model fit the data well and significantly better than the one-factor model (χ² = 59.8 (df = 4), p = 3.141881e-12; AIC = 81.8; SRMR = 0.07; CFI = 0.98; Δχ² = 84.3, Δdf = 1, p = 4.313145e-20). The standardized loadings of cigarette, e-nicotine, and smokeless tobacco use on the first factor were still all over 0.9, while the standardized loadings of cigar and hookah use on the second factor were 1.03 and 0.74, and the two factors were correlated at 0.64 (SE = 0.04; **Figure 3C**). To formally test whether these factors could be considered indistinguishable, we fit a constrained model in which the factor correlation was fixed to 1. This constraint significantly degraded model fit compared to the unconstrained two-factor model (Δχ² = −0.0001, Δdf = 1, p = 1), yielding a model fit nearly identical to the common factor model.

### Genetic correlations between nicotine use and other phenotypes

≥100 Cigarette, e-nicotine, and smokeless tobacco use showed the strongest genetic correlations with psychiatric disorders risk, while cigar and hookah use had significantly weaker genetic correlations with psychiatric phenotypes (**Figure 4**). Across most other substance use traits, cigars and hookah generally showed weaker genetic correlations compared to ≥100 cigarette, e-nicotine, and smokeless use; the main exceptions were cannabis ever-use and drinks-per-week, where cigars and hookah showed relatively stronger correlations. Hookah use exhibited a positive genetic correlation with educational attainment (r_g_ = 0.28, SE = 0.044) and cannabis ever-use (r_g_ = 0.99, SE = 0.078), alongside a negative genetic correlation with BMI (r_g_= −0.16, SE = 0.043), while ≥100 cigarettes, e-nicotine, and smokeless tobacco use showed genetic correlations in the opposite direction for educational attainment and BMI.

**Figure 4.**
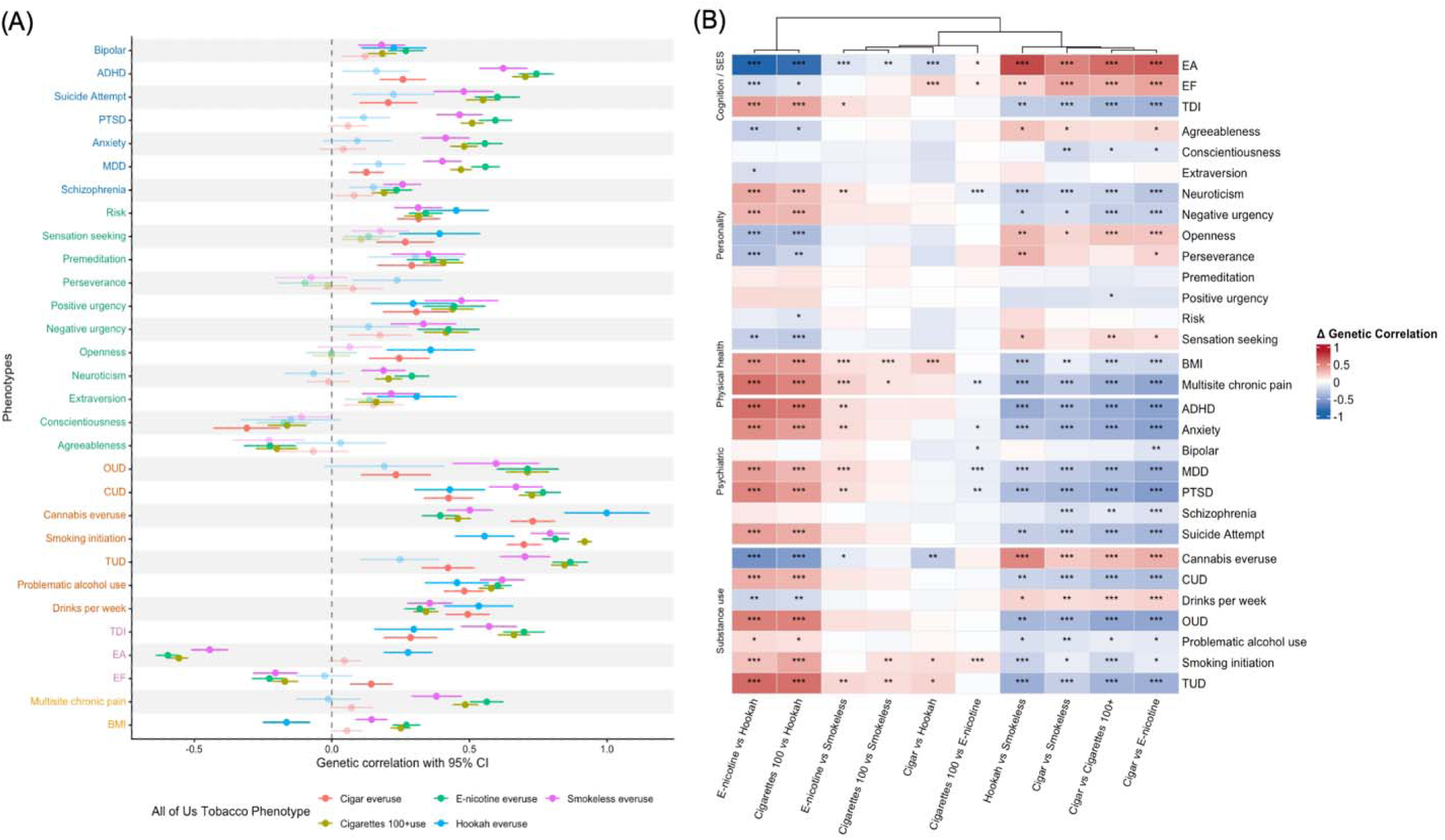
(A) Genetic correlations between nicotine use phenotypes and other traits of interest. (B) Comparisons of genetic correlations across different pairs of nicotine use phenotypes. ADHD = attention deficit hyperactive disorder; MDD = major depressive disorder; PTSD = post-traumatic stress disorder; Risk = risk tolerance; TUD = tobacco use disorder; CUD = cannabis use disorder; OUD = opioid use disorder; TDI = Townsend deprivation index; EA = educational attainment; EF = executive function; BMI = body mass index. The colors of the y-axis labels indicate categories of external traits, and transparency reflects whether the association remains significant after Bonferroni correction. *** represents FDR-adjusted p-value < 0.001; ** represents FDR-adjusted p-value < 0.01; * represents FDR-adjusted p-value < 0.05. Blank squares mean that there was no significant difference in genetic correlations between the two tobacco use phenotypes.

Positive urgency (r_g_ s = 0.30 to 0.47) and risk tolerance (r_g_ s = 0.31 to 0.45) were positively genetically correlated with all nicotine use phenotypes. Negative urgency and neuroticism showed positive correlations only with ≥100 cigarette, e-nicotine, and smokeless tobacco use (r_g_ s = 0.19 to 0.42), while agreeableness showed negative genetic correlations with these same phenotypes (r_g_ s = −0.20 to −0.22); none of these 3 personality traits were significantly associated with hookah or cigar use. Sensation-seeking and openness were positively genetically correlated with cigar and hookah use (r_g_ = 0.24 to 0.39), but showed negligible correlations with ≥100 cigarette, e-nicotine, or smokeless tobacco use. For the remaining two Big Five traits, conscientiousness was negatively genetically correlated across all nicotine use phenotypes, but only statistically significant for ≥100 cigarette (r_g_ = −0.16, SE = 0.035) and cigar use (r_g_ = −0.31, SE = 0.061); extraversion was positively genetically correlated with ≥100 cigarette, hookah use, and smokeless tobacco (r_g_ = 0.16 to 0.31), but was not significantly correlated with e-nicotine or cigar use.

## DISCUSSION

We present the first biobank-scale phenotypic and genetic analysis of different nicotine use modes. In addition to ancestry-specific GWAS conducted in three genetic ancestry groups in *All of Us*, we performed cross-ancestry meta-analysis to increase power for association discovery, resulting in a total of 93 loci identified (87 for ≥100 cigarette smoking, 1 for cigar ever-use, 4 for e-nicotine ever-use, and 1 for smokeless tobacco ever-use). Our study reveals substantial differences in phenotypic distributions across age and sex, and genetic correlations with other health and behavior-related traits.

At the molecular level, gene-based analyses further support the involvement of pathways in cigarette and e-nicotine use related to addiction and neuropsychiatric disorders, as several identified signals map to genes previously implicated in related phenotypes. For example, *THSD7B*, which reached significance in the meta-analysis of ≥100 cigarette and e-nicotine, as well as in the EUR ancestry GWAS of e-nicotine use, was identified as a candidate in genome-wide meta-analysis of nicotine dependence and alcohol dependence^25,26^. Similarly, *NCAM1*, which was significant in analyses of cigarette and e-nicotine use, has been reported to be genetically associated with neurodevelopmental and psychiatric disorders (e.g., bipolar disorder, schizophrenia)^27^. Prior work also showed that genetic variation in this genomic region may influence nicotine dependence indirectly through behavioral mechanisms, such as automatic smoking rituals^28^.

Our genetic correlation analyses showed that hookah and cigar use were strongly genetically correlated with each other (r_g_ = 0.87, SE = 0.08), whereas smokeless tobacco, e-nicotine, and ≥100 cigarette use formed another cluster with strong mutual genetic correlations (r_g_=0.82 (0.047) to 0.92 (0.02)). To further characterize this structure, we applied GenomicSEM to evaluate how these phenotypes load onto shared genetic latent factors. The two-factor model provided a better fit than the common factor model, with cigarette, e-nicotine, and smokeless tobacco use loading strongly on the first factor, while cigar and hookah use loaded primarily on another factor. The two latent factors remained moderately correlated (r_g_ = 0.64, SE = 0.045), suggesting partially shared but distinct genetic influences underlie different nicotine use behaviors. Furthermore, all forms of nicotine use, except cigar smoking, exhibited significant residual variance, suggesting that each mode retains specific genetic variance beyond shared genetic risk. Our results showed important genetic heterogeneity in nicotine use that is often not accounted for in existing GWAS of tobacco use and TUD, which have focused on cigarette smoking.

Genetic correlations with other traits of interest suggested that nicotine use modes may reflect distinct behavioral profiles. ≥100 Cigarette, e-nicotine, and smokeless tobacco use consistently showed positive genetic correlations with psychiatric disorders, negative urgency, and neuroticism. Conversely, cigar and hookah use showed weak or null genetic correlations with psychiatric traits but were positively correlated with openness and sensation-seeking. Unlike cigarettes and e-nicotine, which are packaged for high-frequency, habitual use, cigars and hookah are often used occasionally for social experiences, aligning with their genetic correlation patterns. Our findings are also consistent with prior studies related smoking preference (cigar vs. cigarette) to personality: agreeableness was negatively associated with cigarette smoking but showed negligible associations with cigar use^8^. Notably, hookah use showed a unique positive genetic correlation with educational attainment. This pattern may reflect the social nature of hookah use: hookah smoking is often practiced in group settings^6^ and has been facilitated by the frequent presence of waterpipe establishments near college campuses^7^, which may promote its use as a social activity among young adults. These findings suggest that observed genetic differences across nicotine use modes may partly reflect differences in their social and environmental contexts and patterns of use: cigarette, e-nicotine, and smokeless tobacco use appear more closely related to genetic liability for addiction behaviors and psychiatric traits, whereas cigar and hookah use may reflect more socially driven and occasional use patterns.

Several limitations should be noted. First, the effective sample sizes for e-nicotine, hookah, and smokeless tobacco use in the AFR and AMR subsamples were relatively small (e.g, Neff_AFR,hookah_=33,057, Neff_AMR,hookah_ =36,581; see **Table 1**). These limited sample sizes may have reduced our statistical power to detect genetic effects. We expect future studies with larger and more diverse samples to improve the estimates and thus better characterize the genetic architecture underlying different nicotine use modes across ancestry. Similarly, our genetic correlation analyses were conducted within the EUR subsample. The heritability estimates for most nicotine use phenotypes in AFR (except for ≥100 cigarette use) were insufficiently powered for genetic correlation analyses (z-scores < 4). This limitation may reduce the generalizability of our findings to non-European populations. Second, the case definition for cigarette smoking differed from all other modes: while the assessments for e-nicotine, cigars, hookah, and smokeless tobacco only required ‘ever’ trying the product, the cigarette smoking phenotype required individuals to have smoked at least 100 cigarettes in their lifetime. Because of these different thresholds, our comparative analyses may be confounded by comparing ‘ever’ use for other modes against a higher initiation threshold for cigarettes. For the non-cigarette products, these surveys do not distinguish between experimental use and habitual or dependent use. Consequently, our analyses may reflect genetic influences related to initiation or occasional usage rather than long-term preference or dependence. Future research should extend to phenotypes reflecting regular use and consumption frequency to better characterize genetic contributions to the progression from initiation to addiction. Finally, population-based GWAS like the current study not only capture direct genetic effects but may also reflect gene-environment correlations (i.e., indirect genetic effects) and assortative mating; future within-family genetic analyses will be able to parse direct genetic effects from potential confounders.

Our findings also highlight several additional directions for future research. While we observed substantial phenotypic differences across age and sex groups, we did not explicitly model gene-by-demographic interactions (e.g., gene-by-age or gene-by-sex). Future work should investigate whether genetic influences on nicotine use behaviors differ across age or sex. This consideration may be particularly important for emerging nicotine products such as e-nicotine, which have become widely available only in the past decade. Therefore, patterns of use may differ across birth cohorts, and incorporating cohort-specific analyses may help clarify how genetic liability may interact with changing tobacco environments.

## CONCLUSION

In summary, our study provides comprehensive analyses outlining the shared and distinct genetic architectures underlying five different nicotine use modes. We identified two primary genetic clusters: one consisting of cigarette, e-nicotine, and smokeless tobacco use, and another comprising hookah and cigar use. These clusters exhibit divergent genetic correlation patterns with addiction-related, psychiatric, socioeconomic, and personality traits, suggesting that various nicotine products cannot be treated as a single homogeneous phenotype. Our work potentially helps inform more nuanced public health interventions and clinical strategies tailored to the diverse modes of nicotine use.

## Data Availability

All data is available online to registered users at the All of Us Researcher Workbench.

## Acknowledgments

We would like to thank the research participants and employees of 23andMe Research Institute for making this work possible.

The All of Us Research Program is supported by the National Institutes of Health, Office of the Director: Regional Medical Centers: 1 OT2 OD026549; 1 OT2 OD026554; 1 OT2 OD026557; 1 OT2 OD026556; 1 OT2 OD026550; 1 OT2 OD 026552; 1 OT2 OD026553; 1 OT2 OD026548; 1 OT2 OD026551; 1 OT2 OD026555; IAA #: AOD 16037; Federally Qualified Health Centers: HHSN 263201600085 U; Data and Research Center: 5 U2C OD023196; Biobank: 1 U24 OD023121; The Participant Center: U24 OD023176; Participant Technology Systems Center: 1 U24 OD023163; Communications and Engagement: 3 OT2 OD023205; 3 OT2 OD023206; and Community Partners: 1 OT2 OD025277; 3 OT2 OD025315; 1 OT2 OD025337; 1 OT2 OD025276. In addition, the All of Us Research Program would not be possible without the partnership of its participants. This manuscript is the result of funding in whole or in part by the National Institutes of Health (NIH). It is subject to the NIH Public Access Policy. Through acceptance of this federal funding, NIH has been given a right to make this manuscript publicly available in PubMed Central upon the Official Date of Publication, as defined by NIH.

This work utilized the High Throughput Computing Facility at the Center for Genome Sciences and Systems Biology at Washington University in St. Louis.

ECJ is supported by K01DA051759. PNRV is supported by R90NR021799.

